# Adverse events following immunization during COVID-19 mass vaccination campaigns in the Democratic Republic of Congo: Findings from active safety surveillance

**DOI:** 10.1101/2024.08.18.24312190

**Authors:** Kizito Kayumba, Alemayehu Duga, Mosoka Papa Fallah, Claire Tshidibi, Nicky Lubaki, Barnabas Coulibaly, Benjamin Djoudalbaye, Efire Nora Sylvana, Elizabeth Gonese, Senga Lucy Sembuche, Tedi Tilahun, Murtala Jibril, Carlos Kilowe, Aminu Kuba, Antoine Mafwila Lusala, Nebiyu Dereje, Nicaise Ndembi, Tajudeen Raji, Ngashi Ngongo

## Abstract

**Introduction:** Post-marketing safety surveillance is indispensable for ensuring safety and public trust in vaccines, particularly in African settings where evidence of COVID–19 vaccine safety is less documented. We aimed to identify the types of adverse events following immunization (AEFIs), the overall incidence, and factors associated with AEFIs during mass vaccination campaigns in The Democratic Republic of Congo (DRC).

**Methods:** From December 1 to 29, 2023, a prospective safety surveillance study was conducted on 4,766 individuals in Kinshasa Province in DRC. They were surveyed through phone calls from day 1 to 28 following the administration of the COVID-19 vaccine. We calculated AEFI incidence rates by type of vaccine, sex, and age group. We identified factors associated with AEFIs using multivariable logistic regression models, which were expressed by adjusted odds ratio (aOR) and its 95% confidence interval (CI).

**Results:** A total of 4766 participants were included in the study. The median age of the participants was 36 years, with an interquartile range of 27 to 48 years and 2503 (53%) were females. Most of the participants (94.6%) received the Johnson & Johnson (J&J) vaccine, while 256 (5.4%) received the Pfizer vaccine. Nearly a quarter of participants (23.75%, 95% CI: 22.54%-24.99%) reported AEFIs. The most common AEFIs reported were fever (9.61%, 95%CI: 8.88%-10.48%), injection site pain (9.00%, 95%CI: 8.20%-9.85%), headache (4.11%, 95%CI: 3.57%-4.72%), stiffness (1.51%, 95%CI: 1.18%-1.89 %) and myalgia (1.15%, 95%CI: 0.87%-1.49%). The incidence of AEFIs was higher for the Pfizer vaccine at 34.48% (95% CI: 28.57%-40.54%) compared to 23.15% (95% CI: 21.92%-24.41%) for the J&J vaccine. Participants aged 36 years and above were associated with increased odds of reporting any adverse event (aOR=1.17, 95%CI: 1.02-1.33), injection site pain (aOR=1.36, 95% CI: 1.11-1.66), and two or more signs (aOR=1.46, 95%CI: 1.10-1.94), compared to those below 36 years of age.

**Conclusions:** AEFI reported by about a quarter of participants and its association with vaccine type and older age underscores the need for systematic vaccine safety monitoring in the population. This is critical for developing future vaccination strategies tailored to individuals more susceptible to AEFIs.

## Introduction

Vaccines approved for use in national immunization programmes (NIPs) are considered safe and efficacious based on verifiable evidence from randomized controlled clinical trials (RCTs)^1^. These trials provide robust evidence supporting their safety and effectiveness. However, despite the rigorous evaluation during clinical development, it is important to note that rare side effects may not be detected in the early phases of clinical trials. Post-marketing safety surveillance after introducing the COVID-19 vaccine into public immunization programs must identify rare and previously undocumented adverse events (AEs) to address this. For instance, the non-replicating vector COVID-19 vaccines such as AstraZeneca and Johnson & Johnson have been associated with central venous sinus and other large vessel thrombosis (∼4 per million vaccinations) and thrombocytopenia. ^2^ Similarly, messenger RNA COVID-19 vaccines have also observed rare side effects, such as Pfizer-BioNTech. These include anaphylaxis and myocarditis.^3^ Moreover, the African population was not adequately represented in RTCs. Therefore, the safety of the vaccines can only be determined by post-market safety surveillance.

Given that vaccines are often recommended for healthy individuals, the key to the success of NIPs is public trust in vaccine safety. ^4^ Thus, systematic vaccine safety surveillance is indispensable for ensuring the safety and public trust in vaccines. Safety surveillance is expected to provide additional information on incidence, distribution, and risk factors for expected and unexpected serious and minor AEFIs.^5^ According to the World Health Organization (WHO), all adverse events(AEs) that concern the caregiver and have associated costs and effects should be reported regardless of severity.^6^ The present study aimed to investigate the occurrence of AEFI during COVID-19 mass vaccination campaigns in the Democratic Republic of Congo. By conducting a thorough analysis of AEFI data collected from these campaigns, this study sought to provide valuable insights into the safety profile of vaccines administered. It contributed to the evidence base for future vaccination efforts.

## Methods

### Study Setting and Design

We conducted a prospective follow-up study using an active safety surveillance approach among COVID-19 vaccine recipients in the Democratic Republic of Congo (DRC). As of June 2023, 17.05 million individuals in the DRC have received at least one dose of COVID-19 vaccine.^7^ The Ministry of Public Health of the DRC, through the Expanded Programme on Immunization (EPI), with the support of Africa Centres for Disease Control and Prevention (Africa CDC) and implementing partners, conducted a two-week Rapid Results Initiative (RRI) COVID-19 mass vaccination campaign. From December 1 to 29, 2023 in Kinshasa province, selected vaccine recipients were recruited for the follow-up. The campaign’s primary objective was to accelerate the COVID-19 vaccination efforts and improve the low vaccination coverage in DRC. Africa CDC and EPI conducted COVID-19 vaccine safety surveillance to document AEFI during the campaign.

During this mass vaccination campaign in the DRC, two types of COVID-19 vaccines were administered: the Pfizer-BioNTech mRNA vaccine and the Johnson & Johnson (J&J) non-replicating vector vaccine. The Pfizer-BioNTech vaccine utilizes mRNA technology to train the immune system to recognize the spike protein of the coronavirus.^8^On the other hand, the J&J vaccine delivers the virus’ DNA to host cells using an adenovirus as a delivery vehicle).^9^

### Study participants

The COVID-19 mass vaccination campaign occurred in 159 vaccination sites distributed in 29 health zones of Kinshasa Province. In each vaccination site, 5% of the people vaccinated were randomly selected for safety surveillance, according to the WHO recommendation^5^. Those participants who received COVID-19 vaccines during the campaign consented to participate in the study and provided a reachable phone number.

### Data Collection and Management

We used a structured questionnaire adapted from the WHO safety surveillance tool^5^. The tool captured data on population characteristics such as patient ID, province, profession, sex, and age, as well as vaccine-related information such as batch number, number of doses, type of vaccine, vaccination date, and the record of AEF. The data concerning AEFI included the date of the caller, the date of onset of the event and the type of AEFI. Follow-up calls were made on days 1, 3, 5, 7, 14, 21, and 28 after vaccine administration and data collection forms were used to record information. The AEFI was considered serious in case of an event that results in death, hospitalization, or prolongation of an existing hospitalization, persistent or significant disability or incapacity, congenital anomaly/birth defect, or is life-threatening or is a medically important event or reaction.^5^ Africa CDC recruited and trained a local team of AEFI focal points to conduct follow-up calls to identify and monitor the AEFI. A data collection line listing form with built-in validation rules was used to ensure data quality during data entry.

### Data Analysis

The study’s primary outcome was the experience of at least one AEFI among those receiving at least one dose of any COVID-19 vaccine used during the vaccination mass campaign. Eight Adverse Events for Special Interest (AESI) conditions pre-defined for surveillance based on WHO recommendation, including fever, vomiting, headache, muscle pain, joint pain, body aches, persistent pain at the injection site, diarrhea, and chills, were followed and monitored. We dichotomized the number of events as 2 or more coded as one (1) and 0–1 coded as zero (0) for our logistic regression model for the number of AEs ^14^. The independent variables considered in this analysis included age (dichotomized as <36 years and ≥36 years), sex, type of COVID-19 vaccines, and the number of COVID-19 vaccine doses received.

Descriptive statistics were used to summarize socio-demographic characteristics and vaccine-related information by frequency and proportion for categorical variables and median with interquartile range (IQR) for numerical variables. The cumulative incidence rate of AEFI was provided by proportion along with its respective 95% confidence interval. The multivariate regression models for statistical analysis using the backward-elimination regression approach included all the above-mentioned independent variables. Multicollinearity was assessed by the variance inflation factor (VIF) and the tolerance test (≥0.1). The VIF below ten was considered acceptable to declare lack or absence of multicollinearity. Variables with collinearity were omitted from the models. The goodness of model was assessed by the Hosmer Lemeshow goodness of fit test (P-value >0.05). Multivariable logistic regression models identified factors associated with experiencing AEFI and expressed by adjusted odds ratio (aOR) and its 95% CI. A p-value < 0.05, together with 95%CI, was used to declare statistical significance. Stata IC version 16 (Stata Corp) was used to analyze the data in this study.

### Ethical considerations

After the vaccine was administered, selected participants were informed about the purpose of this surveillance and were asked to willingly participate in this survey. The consent was obtained verbally. The verbal consent was documented on vaccine recipients’ follow up form and witnessed by the enumerator. Those who consented provided their phone numbers to enable follow-up calls. The parents’ phone numbers were provided for the vaccine recipients under 18. To ensure confidentiality, de-identified data was shared with Africa CDC to analyze the vaccine’s safety profile following immunization. Ethical Approval was obtained from the appropriate Institutional Review Board to conduct this survey and publish the findings from this analysis.

## Results

### Population and Vaccine Characteristics

The analysis included 4766 participants, with a median age of 36 (Interquartile Range [IQR]: 27-48). More than half (53%) of the participants were females, and more than 70% of respondents were in the 20—to 49-year age range.

Most participants (94.4%) received the Johnson & Johnson vaccine, and the remaining 5.6% were vaccinated with the Pfizer vaccine. Those taking their second dose were 0.6%, and 99.4% took their first dose (Table 1).

**Table 1:**
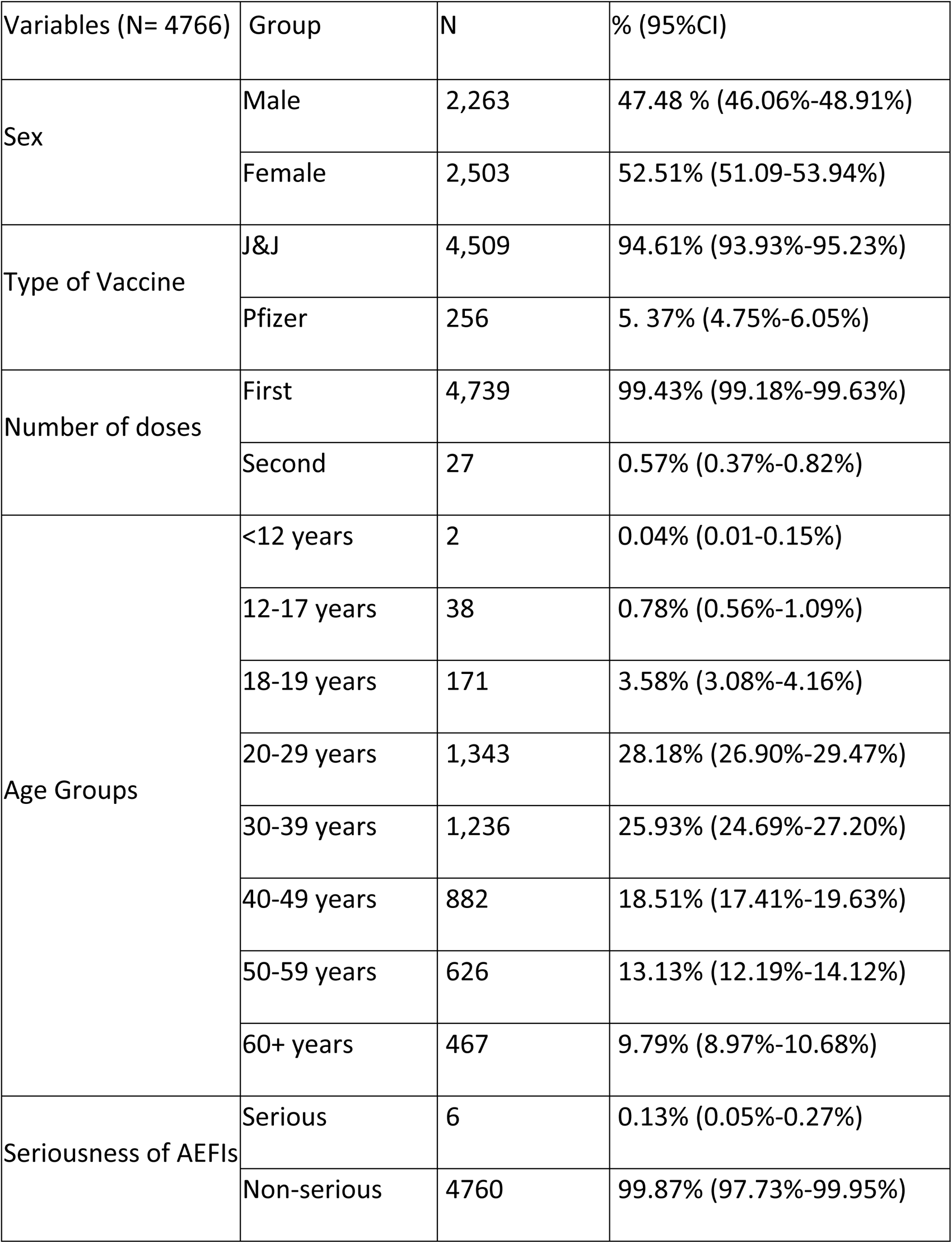
Population and vaccine characteristics.

| Variables (N= 4766) | Group | N | % (95%CI) |
| --- | --- | --- | --- |
| Sex | Male | 2,263 | 47.48 % (46.06%-48.91%) |
|  | Female | 2,503 | 52.51% (51.09-53.94%) |
| Type of Vaccine | J&J | 4,509 | 94.61% (93.93%-95.23%) |
|  | Pfizer | 256 | 5.37% (4.75%-6.05%) |
| Number of doses | First | 4,739 | 99.43% (99.18%-99.63%) |
|  | Second | 27 | 0.57% (0.37%-0.82%) |
| Age Groups | <12 years | 2 | 0.04% (0.01-0.15%) |
|  | 12-17 years | 38 | 0.78% (0.56%-1.09%) |
|  | 18-19 years | 171 | 3.58% (3.08%-4.16%) |
|  | 20-29 years | 1,343 | 28.18% (26.90%-29.47%) |
|  | 30-39 years | 1,236 | 25.93% (24.69%-27.20%) |
|  | 40-49 years | 882 | 18.51% (17.41%-19.63%) |
|  | 50-59 years | 626 | 13.13% (12.19%-14.12%) |
|  | 60+ years | 467 | 9.79% (8.97%-10.68%) |
| Seriousness of AEFIs | Serious | 6 | 0.13% (0.05%-0.27%) |
|  | Non-serious | 4760 | 99.87% (97.73%-99.95%) |

### Incidence and Forms of AEFI Reported

Overall, 1131 (23.75%, 95% CI: 22.55% - 24.99%) reported having developed at least one AEFI during the study period. The most frequent forms of AEFI reported (ranging between 1% to 10%) are fever 458(9.61%, 95%CI: 8.88%-10.48%) followed by pain on injection site 429 (9.00%, 95%CI: 8.20%-9.85%), headache 196 (4.11%, 95%CI: 3.57%-4.72%), stiffness 72 (1.51%, 95%CI: 1,18%-1.89%) and myalgia 55(1.15%, 95%CI: 0.87%-1.49%). The less frequent forms (ranging between 0.1% to 1%) include vomiting 24 (0.50%, 95%CI: 0.32%-0.75%), arthralgia 18(0.38%, 95%CI: 0.22%-0.59%), diarrhea 12(0.25%, 0.13%-0.44%), vertigo 12(0.25%.95%CI: 0.13%-0.44%) and chills 10 (0.21%, 95%CI: 0.10%-0.38%). The unusual AEFIs reported (ranging between 0.01% to 0.1%) are heaviness of arm 4 (0.08%, 95%CI: 0.02%-0.21%), gastritis 2 (0.04%, 95%CI: 0.010%-15%), blood pressure 1(0.02%, 95%CI: 0.01%-0.11%)), blurred vision 1 (0.02%, 95%CI: 0.01%-0.11%), burning sensation 1 (0.02%, 95%CI: 0.01%-0.11%) (Figure 1)

**Figure 1:**
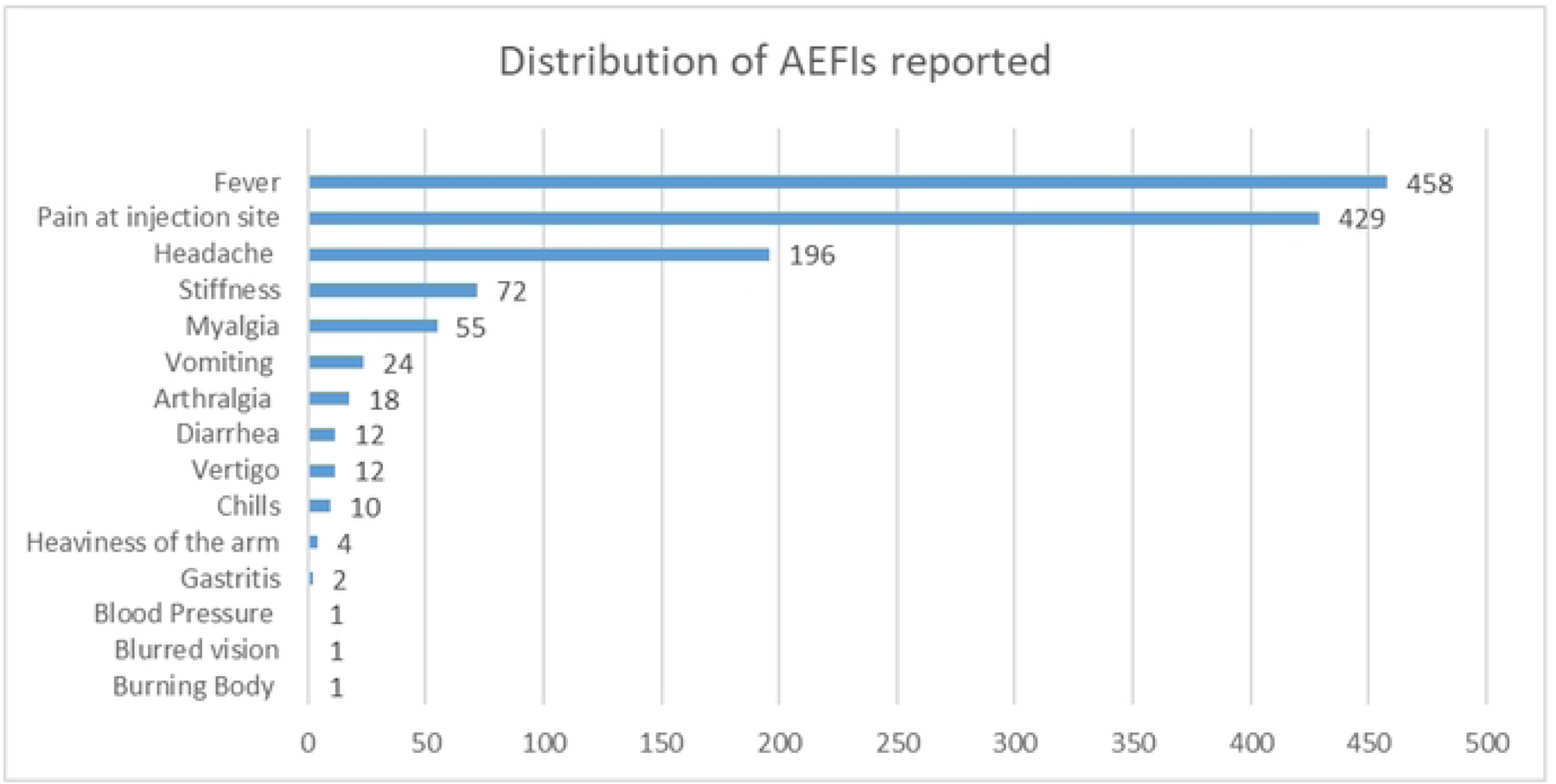
Frequency of reported AEFis

### Factors Associated with the Incidence of AEFIs

In the multivariable analysis, the occurrence of an AEFI was statistically associated with age and the type of vaccine administered. Incidence of AEFI was higher among Pfizer vaccine recipients (34.38%, 95% CI: 28.57%%-40.54%) versus J&J vaccine recipients (23.15%, 95% CI: 21.92%-24.41%); slightly higher among vaccine recipients aged 36 years and more (25.11%, 95%CI: 23.39%-26.88%) versus those below 36 years of age (22.34%, 95% CI: 20.66%-24.08%). The odds of any AEs were about two times higher in the Pfizer vaccine (aOR, 1.73; 95% CI: 1.31-2.28); p<0.001) as compared to J & J vaccine recipients and 17% higher among the older age group (aOR, 1.17, 95%CI: 1.02-1.33; p<0.022) at 36 years and more vs. <36 years. No statistical association was found for gender and number of doses (**Table 2**).

**Table 2:** Factors associated with the incidence of AEFIs.

| Independent Variables | Group | AEFI (-) | AEFI (+) | % AEFI | aOR (95%CI) | P-value |
| --- | --- | --- | --- | --- | --- | --- |
| Gender | Male | 1,908 | 595 | 23.77% | 1.00 (0.87-1.14) | 0.986 |
|  | Female | 1,726 | 537 | 23.70% | 1 (Ref) |  |
| Age | 36 Years and + | 1,819 | 610 | 25.11% | 1.17 (1.02-1.34) | 0.022* |
|  | <36 years | 1,815 | 522 | 22.34% | 1 (Ref) |  |
| Type of Vaccine | Pfizer | 168 | 88 | 34.38% | 1.73 (1.32-2.28) | 0.001* |
|  | Johnson & Johnson | 3,466 | 1,044 | 23.15% | 1 (Ref) |  |
| Number of doses administered | 1<sup>st</sup> dose | 3,613 | 1,126 | 23.76% | 1.87(0.73-4.79) | 0.190 |
|  | 2<sup>nd</sup> dose | 21 | 6 | 22.22% | 1 (Ref) |  |

\* Significant factors, p<0.05

Fever was higher for Pfizer vaccine recipients (12.50%, 95%CI; 8.71%-17.18%) than J&J vaccine recipients (9.45%, 95%CI: 4.35%-10.34%). Recipients of the Pfizer vaccine were more likely susceptible to developing fever than recipients of the J&J vaccine (aOR=1.47, 95%CI: 1,02-2.16; p<0.048) (**Table 3**).

**Table 3:** Factors Associated with Fever.

| Independent Variables | Group | Fever(-) | Fever (+) | % Fever | aOR (95%CI) | P-value |
| --- | --- | --- | --- | --- | --- | --- |
| Gender | Male | 2,255 | 248 | 9.91% | 1.07 (0.89-1.30) | 0.452 |
|  | Female | 2,053 | 210 | 9.28% | 1 (Ref) |  |
| Age | 36 years and + | 2,196 | 233 | 9.59% | 0.96(0.82-1.20) | 0.963 |
|  | <36 years | 2,112 | 225 | 9.63% | 1 (Ref) |  |
| Type of Vaccine | Pfizer | 224 | 32 | 12.5% | 1.47(1,002-2.16) | 0.048* |
|  | Johnson & Johnson | 4,084 | 426 | 9.45% | 1 (Ref) |  |

\* Significant factors, $p < 0.05$

The proportion of persistent pain at the injection site was higher for Pfizer vaccine recipients (15. 23%, 95%CI: 11.06%-20.23%%) vs J&J vaccine recipients (8.65%, 95%CI: 4.29%-9.51%). Respondents vaccinated with the Pfizer Vaccine had a 76% increased risk of reporting pain at the injection site than those vaccinated with the J&J vaccine (aOR=1.76,95%CI: 1.21-5.56; p<0.003). Respondents aged 36 years and older also had a 36% increased risk of reporting pain at the site of injection compared to those aged below 36 years (aOR=1.36, 95%CI: 1.11-1.66; p<0.003). Although not statistically significant, the incidence of persistent pain at the injection site was higher after the 2nd dose (18.52%, 95%CI 6.30%-38.08%) than the first dose (8.95%, 95%CI: 8.15-%-9.79%) (**Table 4**)

**Table 4:** Factors associated with persistent pain at the injection site.

| Independent Variables | Groups | Pain (-) | Pain (+) | % Injection site pain | aOR (95%CI) | P-Value |
| --- | --- | --- | --- | --- | --- | --- |
| Gender | Male | 2,280 | 223 | 8.91% | 0.97(0.79-1.18) | 0.756 |
|  | Female | 2,057 | 206 | 9.10% | 1 (Ref) |  |
| Age group | 36 Years and + | 2,181 | 248 | 10.21% | 1.36(1.11-1.66) | 0.003* |
|  | <36 years | 2,156 | 181 | 7.74% | 1 (Ref) |  |
| Type of Vaccine | Pfizer | 217 | 39 | 15.23% | 1.76(1.21-5.56) | 0.003* |
|  | Johnson & Johnson | 4,120 | 390 | 8.65% | 1 (Ref) |  |
| Number of doses administered | 1<sup>st</sup> dose | 4,315 | 424 | 8.95% | 0.75(0.27-2.12) | 0.598 |
|  | 2<sup>nd</sup> dose | 22 | 5 | 18.52% | 1 (Ref) |  |

\* Significant factors, $p < 0.05$

Headache was more reported among Pfizer vaccine recipients (9.77%, 95%CI: 6.42%-14.07) than J&J vaccine recipients (3.79%, 95%CI: 3.25%-4.39%), muscle pain was reported by 3.52% (95%CI: 1.62%-6.57%) of Pfizer vaccine recipients vs 1.02% (95%CI; 0.75%-1.35) of J&J vaccine recipients. The more significant proportion of Pfizer vaccine recipients reported two or more symptoms (10.94%, 95%CI: 7.39%-15.41%) than J&J vaccine recipients (4.06%, 95%CI: 3.50%-4.65%). Respondents aged 36 years and more had a 46% increased risk of complaining of 2 side effects or more compared to those aged below 36 years (aOR=1.46, 95%CI: 1.10-1.94; p<0.008).

While all AEs were more predominant in Pfizer than in J&J vaccine, the odds were particularly higher for headache (aOR=2.8, 95%CI: 1.80-4.42; p<0.001) (Table 5), myalgia (aOR=3.79, 95%CI: 1.83-7.84; p<0.001) (Table 6) and reporting two signs or more (aOR=3.02, 95%CI: 1.97-4.63; p<0.001). (Table 7)

**Table 5:** Factors associated with headache.

| Independent Variables | Groups | Headache (-) | Headache (+) | % Headache | OR (95%CI) | P-Value |
| --- | --- | --- | --- | --- | --- | --- |
| Gender | Male | 2,398 | 105 | 4.19% | 1.05 (0.79-1.40) | 0.707 |
|  | Female | 2,172 | 91 | 4.02% | 1 (Ref) |  |
| Age | 36 Years and + | 2,329 | 100 | 4.12% | 1.00 (0.75-1.34) | 0.970 |
|  | <36 years | 2,241 | 96 | 4.11% | 1 (Ref) |  |
| Type of Vaccine | Pfizer | 231 | 25 | 9.77% | 2.82 (1.80-4.42) | 0.001* |
|  | Johnson & Johnson | 4,339 | 171 | 3.79% | 1 (Ref) |  |
| Number of doses administered | 1<sup>st</sup> dose | 4,544 | 190 | 4.11% | 2.85(0.37-21.93) | 0.313 |
|  | 2<sup>nd</sup> dose | 21 | 6 | 3.70% | 1 (Ref) |  |

\* Significant factors, p<0.05

**Table 6:**
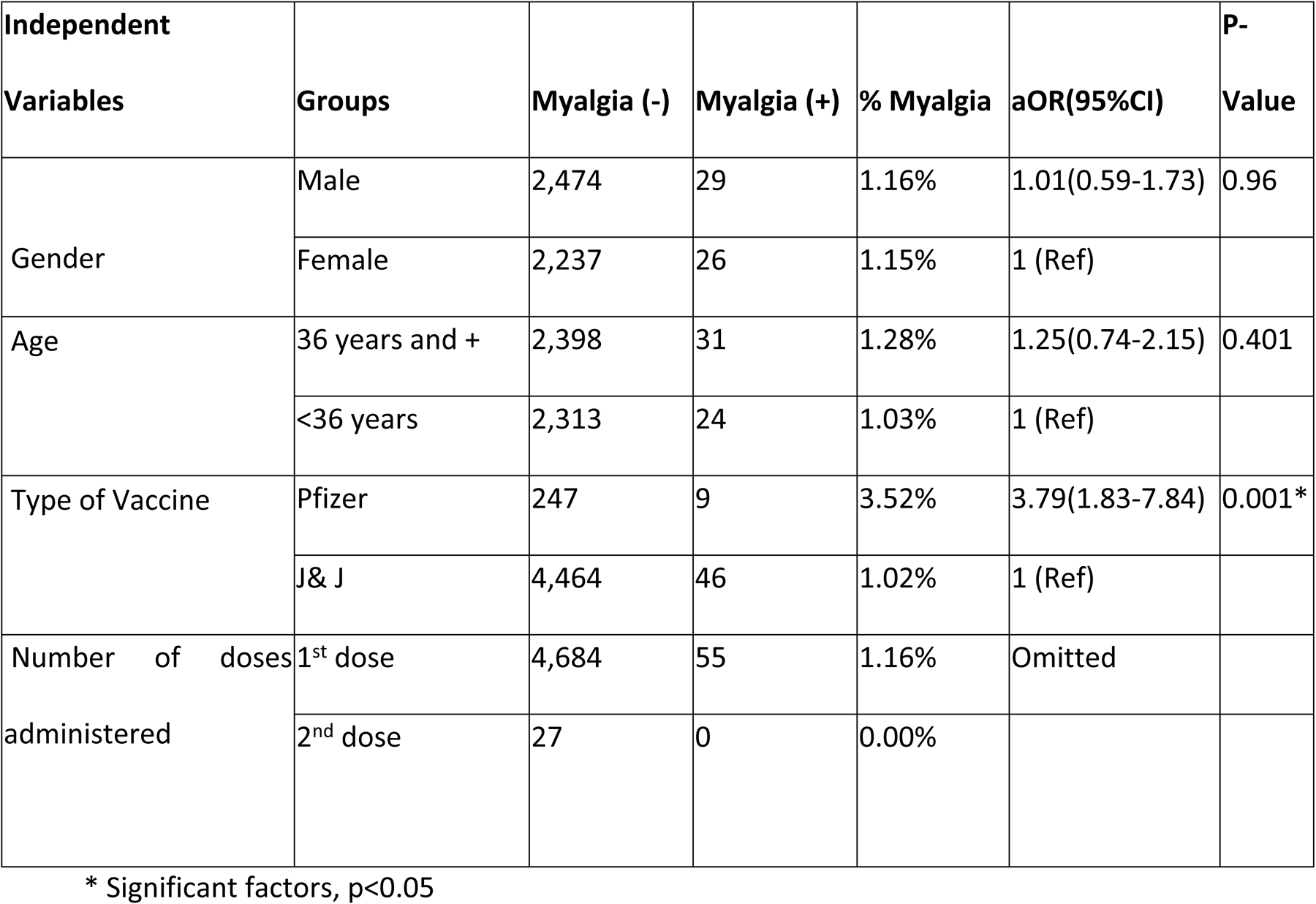
Factors associated with muscle pain.

| Independent Variables | Groups | Myalgia (-) | Myalgia (+) | % Myalgia | aOR(95%CI) | P-Value |
| --- | --- | --- | --- | --- | --- | --- |
| Gender | Male | 2,474 | 29 | 1.16% | 1.01(0.59-1.73) | 0.96 |
|  | Female | 2,237 | 26 | 1.15% | 1 (Ref) |  |
| Age | 36 years and + | 2,398 | 31 | 1.28% | 1.25(0.74-2.15) | 0.401 |
|  | <36 years | 2,313 | 24 | 1.03% | 1 (Ref) |  |
| Type of Vaccine | Pfizer | 247 | 9 | 3.52% | 3.79(1.83-7.84) | 0.001* |
|  | J& J | 4,464 | 46 | 1.02% | 1 (Ref) |  |
| Number of doses administered | 1<sup>st</sup> dose | 4,684 | 55 | 1.16% | Omitted |  |
|  | 2<sup>nd</sup> dose | 27 | 0 | 0.00% |  |  |

\* Significant factors, p<0.05

**Table 7:** Factors associated with the number of signs reported by an individual.

| Independent Variables | Groups | < 2 signs | 2 signs and + | % 2 signs and + | aOR(95%CI) | P-value |
| --- | --- | --- | --- | --- | --- | --- |
| Gender | Female | 2,163 | 100 | 4.43% | 0.99 (0.76-1.32) | 0.992 |
|  | Male | 2,392 | 111 | 4.42% | 1 (Ref) |  |
| Age | 36 Years and + | 2,303 | 126 | 5.19% | 1.46 (1.10-1.94) | 0.008* |
|  | <36 years | 2,252 | 85 | 3.64% | 1 (Ref) |  |
| Type of Vaccine | Pfizer | 228 | 28 | 10.94% | 3.02 (1.97-4.63) | 0.001* |
|  | Johnson & Johnson | 4,327 | 183 | 4.06% | 1 (Ref) |  |
| Number of doses administered | 1<sup>st</sup> dose | 4,529 | 211 | 4.43% | 3.37 (0.44-25.83) | 0.242 |
|  | 2<sup>nd</sup> dose | 26 | 1 | 3.57% | 1 (Ref) |  |

\* Significant factors, $p < 0.05$

## Discussion

The clinical trials during the development of COVID-19 vaccines were mainly conducted in developed nations, so they might not optimally represent the safety situation in the African setting. The findings from our safety surveillance analysis indicate that a significant proportion of COVID-19 vaccine recipients experienced AEFIs. Specifically, the study revealed that about a quarter of individuals who received the vaccine reported any AEFIs and the AEFIs were related to the type of vaccines administered and the age of the recipients. This finding is consistent with clinical trials. In the results from the clinical trial on the safety of mRNA vaccines, more mRNA recipients than placebo recipients reported any adverse event (27% and 12%, respectively) or a related adverse event (21% and 5%).^11^ There was a slight difference between the first and second dose, with 23.76% (95%CI: 22.55%-24.99%) experiencing AEFIs after receiving the first dose and 22.22% (95%CI: 8.62%-42.26%) after the second. Furthermore, it was observed that a higher proportion of individuals who received the Pfizer vaccine (34.38%, 95% CI: 28.57%%-40.54%) reported AEFIs compared to those who received the J&J vaccine (23.15%, 95% CI: 21.92%-24.41%).

Considerable differences were observed in the trends of AEFIs compared with results from the participant-reporting studies. The UK app study showed that 71.9% (dose 1) and 68.2% (dose 2) reported local side effects. ^12^ In the US online cohort study, AEs were reported to be 64.9% partially vaccinated and 80.3% fully vaccinated with the mRNA vaccine.^13^ The proportion of AEs was generally lower in our study. We can speculate that the difference might be due to potential age differences in the study cohort. In our study, the population mainly comprised of younger individuals (median age of 36 years) compared with the studies mentioned above (54 years; UK study; 59−64 years; US study), contrasting trends of AEFIs and association of age was observed, unlike other studies. Moreover, we did not assess whether participants were pre-medicated before vaccination. A study conducted in Ghana revealed that almost half of the respondents pre-medicated with paracetamol before vaccination.^14^ The importance of this factor cannot be ruled out since it may contribute significantly to how an individual’s immune system responds to the vaccine. This creates an avenue for further research to determine if this is the case. Another explanation might be that for participant-reporting studies, the vaccinees without AEs might be less motivated to proactively report Zero AEFI, leading to more reports from those who experienced AEs.

### Forms of AEFI

Findings from this study showed that the most common adverse event experienced among the majority after COVID-19 vaccination included fever (458 cases), pain on injection site (429 cases), headache (196 cases), stiffness (72 cases), and myalgia (55 cases). The adverse events experienced were no different from those experienced across the world.^15^ In the Ethiopian study conducted by Jerso et *al*. on healthcare workers vaccinated against COVID-19^16^, the most prevalent symptoms experienced in descending order include - pain at the injection site (64.1%), fatigue (35.7%), headache (28.9%), joint pain (26.5%) and muscle pain (21.5%). In the study conducted in Ghana, the prevalence of pain at the injection site also hovered around a similar figure of 65.8%, which was also the most common symptom experienced post-COVID-19 vaccination. The second most common symptom was headache (57.5%), followed by the following—tiredness (55.8%), fever (51.7%), chills (39.6%), and muscle pains (38.3%).^14^ In our study, pain at the injection site was the second most common symptom after fever. It is, however, difficult to compare those studies to ours because J&J and Pfizer were not the predominant vaccines administered in those studies. However, the running theme seems to be that pain at the injection site was among the first 2 most common adverse events or side effects in all studies mentioned.

Although not statistically significant, the occurrence of injection site pain in our study was higher among vaccine recipients of the second dose than those with the first dose (18.5% vs 8.9%). These results corroborate the findings of the clinical trials that revealed that the proportion of almost all AEFIs was higher in recipients of the second dose.^17,18^ Similar to previous studies, injection site pain and muscle pain were among the most predominant AEs.^19,20^

### Seriousness of AEFIs

In our findings, 6 (0.13%) serious AEs were reported. In the clinical trial on the safety of mRNA vaccines, the incidence of serious adverse events was similar in the vaccine and placebo groups (0.6% and 0.5%, respectively).^17^

### Factors Associated with AEFIs

Our findings revealed that the vaccine type was the most significant factor that may impact AEs. Pfizer vaccine recipients were significantly associated with greater odds of reporting fever (aOR=1.47, 95%CI: 1,02-2.16; p<0.048), headache (aOR=2.8, 95%CI: 1.80-4.42; p<0.001), injection site pain (aOR=1.76,95%CI:1.21-5.56; p<0.003)), and having more than 2 signs (aOR=3.02, 95%CI: 1.97-4.63; p<0.001), compared to the J&J vaccine recipients. Particularly, Pfizer (an mRNA vaccine) was significantly associated with higher odds of AEs than J&J. These results were consistent with the findings of clinical trials^17,18^ survey-based studies^13,21,22^ and a government-sponsored surveillance study^23^.

Of the baseline characteristics, female gender was not one of the evident factors associated with AEFIs in this study. This was considerably different from previous findings, which showed that the overall incidence of AEs in women was generally higher than that in men.^13,21^ A notable finding from these studies was that female participants reported menstrual disorders and unexpected vaginal bleeding after mRNA vaccination. Menstrual disorders and unexpected vaginal bleeding should be carefully monitored in future surveys.

### Limitations

Our results should be interpreted with caution given the following limitations. First, there might be potential misclassification of outcomes, as AEFIs reported by the participants were not medically reviewed and classification of the AEFI has not been conducted. Secondly, our survey focused only on specific AEFIs using a pre-identified list, limiting the scope and potentially excluding other AEFIs. Thirdly, as we only examined the short-term safety of COVID-19 vaccines, further studies are required to evaluate the long-term AEs. Lastly, this survey did not assess for any pre-existing medical conditions or whether participants were pre-medicated before vaccination.

## Conclusion

In conclusion, this survey conducted in Kinshasa (DRC) revealed that approximately 24% of COVID-19 vaccine recipients aged 5−90 years reported AEFIs, and most of AEFIs were mild and transient. Fever, Injection site pain, headache, and muscle pain were the most predominant AEFIs reported by COVID-19 vaccine recipients. We also found that vaccination with the Pfizer vaccine and age of 36 years or more were associated with the incidence of AEFIs. The Pfizer vaccine was significantly associated with greater odds of reporting fever, headache, muscle pain, injection site pain, and more than 2 signs compared to the J&J vaccine. Individuals aged 36 years or more were associated with greater odds of reporting pain at injection site and having more than 2 AEFIs. Further safety surveys monitoring all potential factors are needed to generate more evidence on COVID-19 vaccine safety in an African population. This information will support establishing future vaccination strategies tailored to individuals potentially susceptible to AEFIs.

## Data Availability

The quantitative data analyzed is not publicly available, being the property of the DRC Ministry of Health however, access may be granted with reasonable request and approval by the Ministry

## Acknowledgments

We thank the participants in this study for their great contribution to safety surveillance and the Ministry of Health for sharing data with Africa CDC to enable this analysis.

